# A CROSS-SECTIONAL SURVEY OF THE PREVALENCE AND THE RISK FACTORS OF TOBACCO USE AMONGST CONSTRUCTION ARTISANS IN EKITI STATE, NIGERIA

**DOI:** 10.1101/2021.04.08.21255137

**Authors:** Charles Oluwatemitope Olomofe, Caryl M Benyon, Kabir Adekunle Durowade, Oluwafunmike Ruth Olomofe

## Abstract

**Background:** Construction artisans are those who perform skilled work relating to the erection or assembly of a large structure. These artisans are prone to tobacco use. This study aims to assess the prevalence and the associated risk factors of tobacco use amongst construction artisans in Ekiti State, Nigeria.

**Materials and Methods:** This cross-sectional study employed a multi-stage sampling technique to select participants (carpenters and bricklayers) artisans, journeymen, and their apprentices who were working in Ekiti State. Chi-square and logistic regression were used to test for association in bivariate and multivariate analyses respectively.

**Results:** The prevalence of ever-smoke amongst respondents was 19.3%. Multivariate analysis showed that artisans who were within the age range 31-40 years were four times more likely to use tobacco (OR=3.410; CI=1.476-7.878). Similarly, being in school and divorced/separated were associated with tobacco use.

**Conclusion:** Noting the increased prevalence of tobacco use among construction artisans when compared to the general population, and few self-reported cases of addiction amongst users demands action from communities and government at all levels.

## BACKGROUND

The construction industry plays a major and significant role in the employment creation and economic growth of many nations. As high as eighty percent of people work in the informal sector (not employed by the government) in Nigeria.^1^ and construction workers constitute a major chunk of this population.^1^ The construction industry in Nigeria also contributes significantly to the gross domestic product of the country.^1,2^ Construction artisans (CA) are those who perform skilled work relating to the erection or assembly of large structures such as buildings, roads, and bridges with their hands.^3,4^ The CA is equipped mainly with vocational education; this may be acquired formally or informally through observation, apprenticeship, and short learning cycle.^5^ Occupations such as carpenters, plumbers, electricians, welders, bricklayers, and painters are included in this category.^4^

Tobacco smoking is the leading cause of preventable morbidity and mortality worldwide. Every day over 13 000 people worldwide die from tobacco and nearly 6 million people succumb to tobacco-related illnesses annually.^6^ More so, assuming current patterns of tobacco use and intervention efforts, WHO projects that from 2000 to 2030 the number of smokers will rise from 1.2 billion to 1.6 billion and the annual number of deaths will increase from 4.9 million to 10 million.^6^ Moreover, about 600,000 non-smokers die yearly because of being exposed to secondhand smoke. Research shows that nearly 80% of the world’s 1 billion smokers live in low and middle-income countries (LMICs).^6^ The increasing burden of non-communicable diseases in developing countries with tobacco use being implicated as a risk factor in a number of these diseases is a clear call to make tobacco control a central theme.^7^ Sustainable Development Goal 3 (SDG 3) underscores this by setting a target to strengthen the implementation of the World Health Organization Framework Convention on Tobacco Control (WHOFCTC) in all countries by the year 2030.^8^

The prevalence of smoking amongst CA in Nigeria is 9.6%^3^ and it is greater than 3.9% which is the prevalence of smoking in the general Nigerian population.^9^ CA such as carpenters, plumbers, tillers has higher rates of smoking than workers in other occupations.^10^ In the United States, compared to all other occupational groups, construction workers had the highest number of ever-smokers (48% compared to 39% for all other occupations combined).^11^

This group of artisans works under stressful conditions, and this makes them prone to some behavioral manifestations.^3^ Coupled with their stressful work environment, CA also belongs to the lower socio-economic cadre in Nigeria, people who lack access to information and social amenities.^12^ These socio-demographic and socio-economic inequalities, on the one hand, make CA indulge in the use of tobacco, and on the other hand make them unable to appreciate the health implications of doing so.^12^

According to the health belief model, the perceived susceptibility to illness and personal benefits are critical factors in health-related behavior.^13^ In this light, most CA is not aware of the consequences of smoking and they do not think they are vulnerable to, say, lung cancer. This low perception of illness coupled with the ‘enjoyment’ and increased work speed (benefits) these workers derived from smoking^3^ makes them persist in this unhealthy behavior. Ajzen and Fishbein’s proposition (1980) in the theory of reasoned action also governs the tendency of artisans to indulge in this risky behavior.^13^

There is a paucity of data on the status and wellbeing of the artisan in Nigeria, but as the global economy continues to shrink and unemployment remains high, artisanship may provide a major source of livelihood to the teeming population of unemployed youths. It is therefore imperative to make the overall health status of these workers a priority. This study attempted to determine the prevalence of tobacco use amongst CA and identified the factors responsible for this burden in Ekiti State, Nigeria.

## METHODS

The study was carried out in Ekiti State, which is one of the thirty-six states of Nigeria. It is in the south-western part of the country. The State has 16 Local Government Areas (LGAs), and three senatorial districts. Ekiti State is a culturally homogenous state, inhabited predominantly by the Ekiti sub-ethnic group of the Yorubas. The study population consisted of CA, journeymen, and their apprentices who were carpenters and bricklayers. Research approval was obtained from the University of Liverpool ethics committee and the Ethics and Research Review Committee of the Federal Teaching Hospital, Ido Ekiti, Ekiti State, Nigeria.

The study was conducted using a pre-tested, semi-structured questionnaire and included CA, journeyman, or their apprentice who were above the age of 18 years and consented to participate in the study. The minimum sample size was determined to be 232 at a confidence level of 95% based on the proportion of tobacco use amongst artisans in a previous study 9.6% in a previous study^3^ and a 5% margin of error.

The questionnaire was adapted from the contents of validated World Health Organization/Centre for Disease and Control Global Adult Tobacco Survey (GATS) and the WHO MONICA project protocol for recording smoking history.^9^

It comprised of two sections; Socio-demographic characteristics of respondents and respondents are smoking history. The pre-test of the questionnaire was done on 10% of the subjects in Ido/Osi LGA which was not included in the study. The pretested questionnaires with participants’ information sheets were administered directly to the artisans at their place of work by the researcher. The data was collected from November 2018 to January 2019.

Data collation and editing were conducted manually to detect omissions and to ensure uniform coding. The data was entered into a computer and analysis was completed using SPSS version 21. All categorical variables, frequency tables, and cross-tabulations were generated (Jekel *et al*., 2001). A chi-square test was used to determine the statistical significance and association between dependent and independent variables. A chi-square was preferred because both variables were categorical.^14^ A multivariate analysis using logistic regression was also carried out to determine the factors associated with smoking. A logistic regression analysis was preferred because every smoke is a dichotomous variable (Yes or No) and many independent variables can be tested for association in this analysis.^14^ The level of significance was predetermined at a p-value of less than 0.05 at a 95% confidence level.

## RESULTS

A total of 236 CA were approached in the chosen communities in Ekiti State, four (4) declined to participate citing work schedule and personal engagements as reasons. Among the 232 that responded, only 228 responded well (4 provided incomplete data) giving a 98.3% response rate. Most of the respondents were between the ages of 21-30 years. All respondents were males. Almost half (46.9%) of the respondents had secondary education as their highest level of education, and only 21.1 % had tertiary education. Almost three-fifths (57%) of the respondents were married while the remaining 43% were either single (33.3%), Divorced separated (6.6%), or widowed (3.1%). Two-fifths (39.9%) of the respondents were apprentices but a third (33.8%) of the respondents had more than 20 years of work experience. Over 60% of the respondents were adept in their work while about 40% were an apprentice. A third (33.8%) of the CA had more than 20 years of experience at the job **(Table 1)**.

**Table 1:**
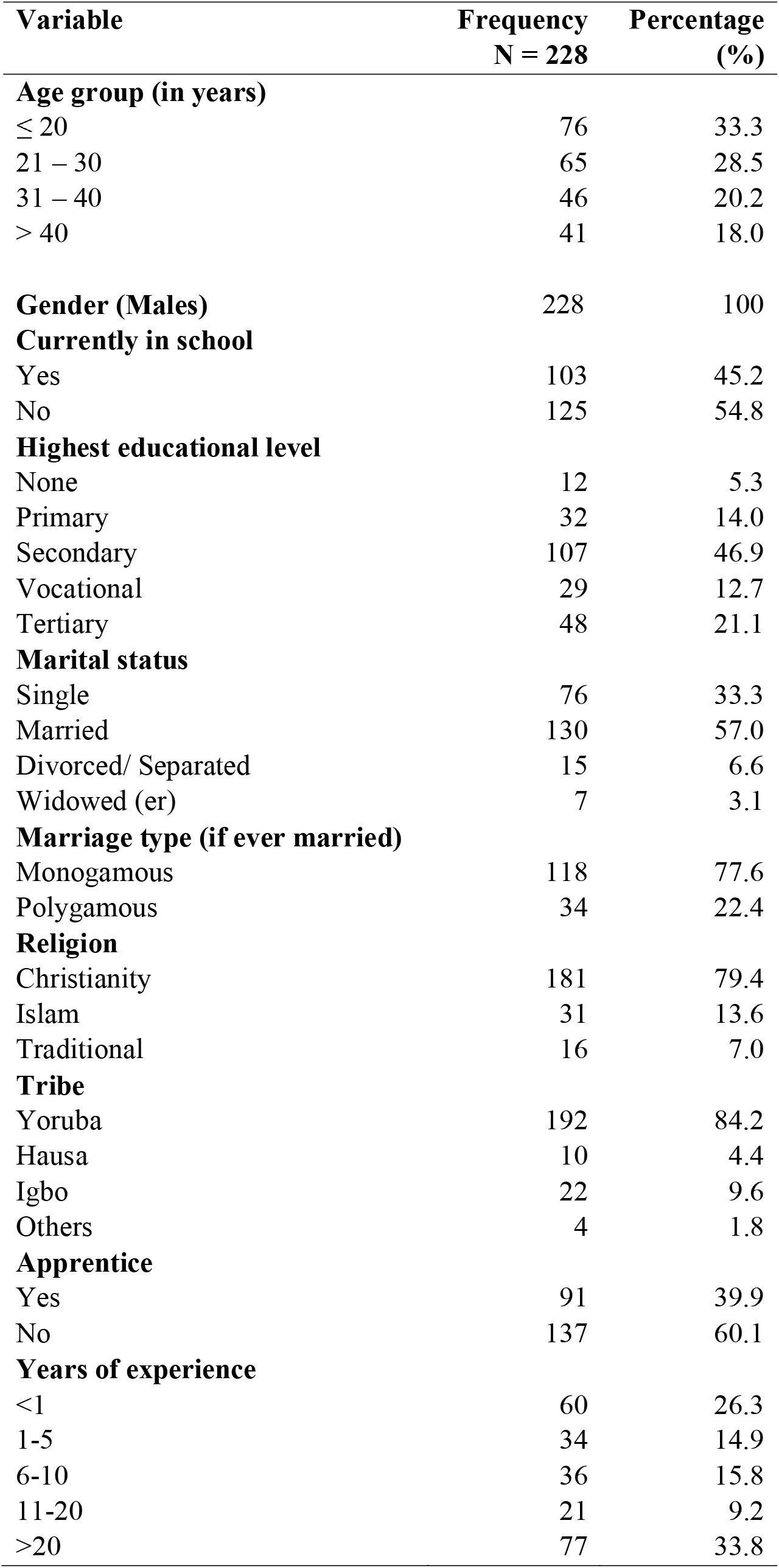
Socio-demographic characteristics of respondent construction artisans.

Amongst the respondents, 19.3% had ever smoked a cigarette in the last 12 months while more than 80% of the CA had not. More than half (53.2%) of ever smokers started smoking before the age of 20 years and two-thirds (65.9%) of the CA admitted to having started the habit because of friends. Half (50%) of the ever-smokers still indulge in the habit, and 90% of them continued in this habit for pleasure they derived from smoking **(Table 2)**. Just 9.1% of the current smokers indulge in the habit for recognition amongst peers. More than two-thirds (68.2%) of the CA smokes between 1-5 cigarette sticks per day, and 31.8% smokes over 5 cigarette sticks per day. Only 31.8% of ever smokers amongst the CA believed that the nature of their job demands they smoke **(Table 2)**. Furthermore, 86.4% admitted to the fact that the smoking habit of their colleagues enticed them to smoke cigarettes. All the respondents were aware that smoking is injurious to health and all had to access the health information on tobacco use. However, 90.9% of the CA who currently smokes were willing to quit but 9.1% did not want to quit smoking because feel they were addicted and could not imagine their life without a cigarette **(Table 2)**.

**Table 2:** The smoking history of construction artisans in this study.

| **Variable** | **Frequency**<br>**N = 44** | **Percentage**<br>**(%)** |
| --- | --- | --- |
| **Ever smoke a cigarette in the last 12 months** |  |  |
| Yes | 44 | 19.3 |
| No | 184 | 80.7 |
| **Total= 228** |  |  |
| **Age at first smoke** |  |  |
| < 20 years | 23 | 52.3 |
| ≥ 20 years | 21 | 47.7 |
| **Reasons you started** |  |  |
| Friends | 29 | 65.9 |
| Adventure | 15 | 34.1 |
| **Currently, smoke** |  |  |
| Yes | 22 | 50.0 |
| No | 22 | 50.0 |
| **Reason for still indulging (n=22)** |  |  |
| Pleasure | 20 | 90.9 |
| Recognition | 2 | 9.1 |
| **Average no of sticks per day (n=22)** |  |  |
| 1 – 5 | 15 | 68.2 |
| > 5 | 7 | 31.8 |
| **Nature of work demands to smoke** |  |  |
| Yes | 14 | 31.8 |
| No | 30 | 68.2 |
| **No of my co-workers smoking** |  |  |
| < 5 | 36 | 81.8 |
| ≥ 5 | 8 | 18.2 |
| **Aware smoking is injurious to your health** |  |  |
| Yes | 42 | 95.5 |
| No | 2 | 4.5 |
| **A co-worker's smoking habit entices you to smoke** |  |  |
| Yes | 38 | 86.4 |
| No | 6 | 13.6 |
| **Want to quit smoking (n=22)** |  |  |
| Yes | 20 | 90.9 |
| No | 2 | 9.1 |

There was a statistically significant association between age and tobacco use amongst CA (p<0.001) **(Table 3)**. CA within the age range 31-40 and those ≤ 20 years have more smokers than those in other age groups. Similarly, there was a statistically significant association (p=0.039) between smoking and being in school **(Table 3)**. CA who were in school smoke more than those who were not in school. There was also a significant association between tobacco use and not being an apprentice (p<0.001). CA who were adept in their occupation smoke more than those who were an apprentice. In a similar vein, there was a statistically significant association between years of experience and tobacco use (p=0.044). CA with over 20 years of experience smoke more than those with fewer years of experience.

**Table 3:**
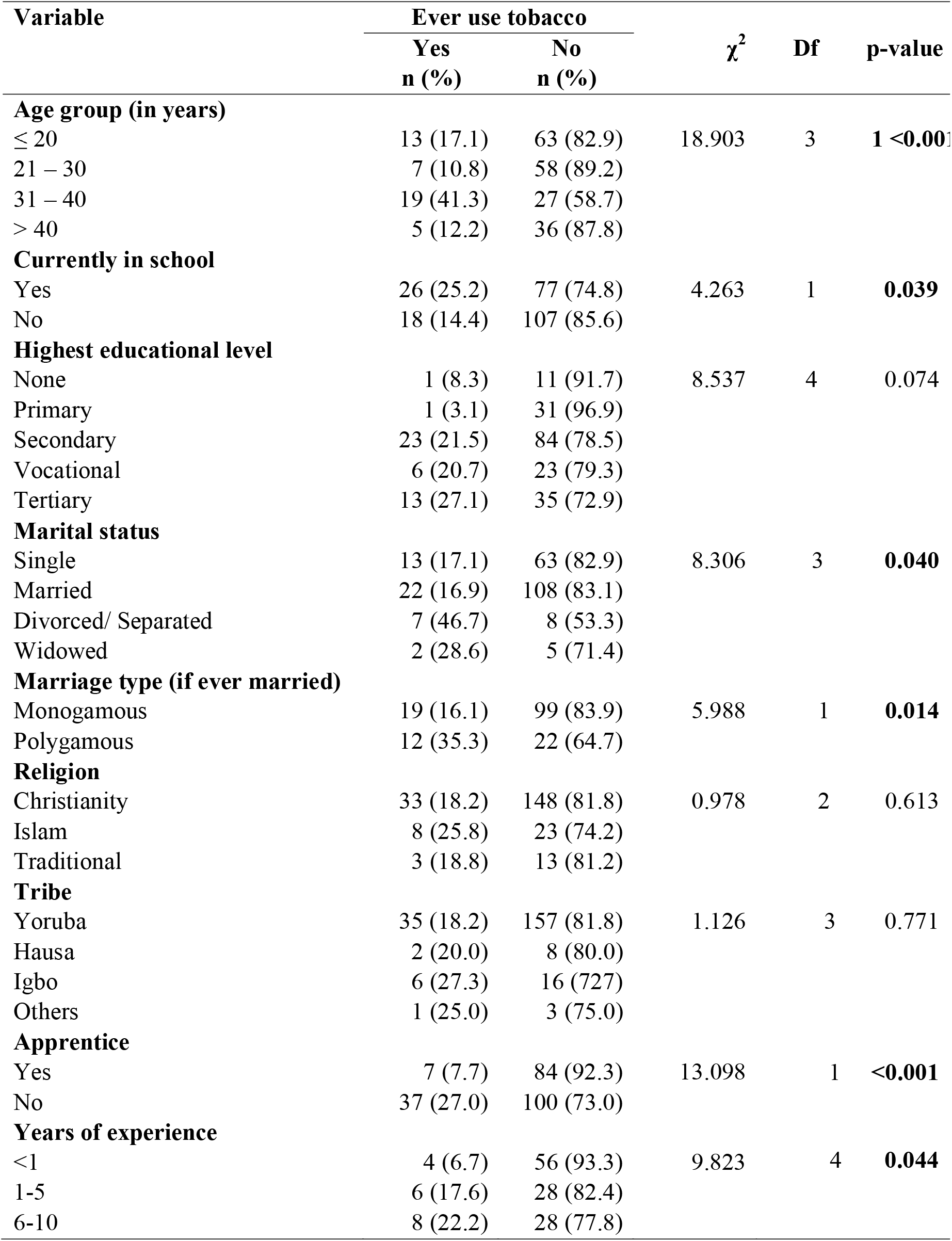
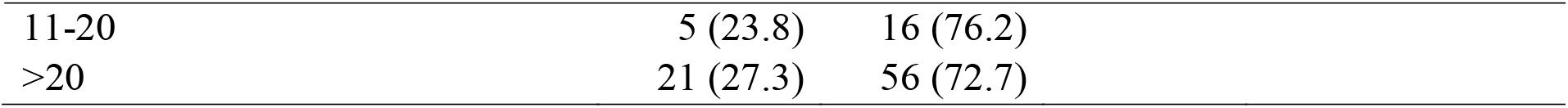
Socio-demographic factors associated with ever smokers among respondent construction artisans.

| Variable | Ever use tobacco | | $\chi^2$ | Df | p-value |
| --- | --- | --- | --- | --- | --- |
|  | Yes<br>n (%) | No<br>n (%) |  |  |  |
| **Age group (in years)** |  |  |  |  |  |
| ≤ 20 | 13 (17.1) | 63 (82.9) | 18.903 | 3 | **1 <0.001** |
| 21 – 30 | 7 (10.8) | 58 (89.2) |  |  |  |
| 31 – 40 | 19 (41.3) | 27 (58.7) |  |  |  |
| > 40 | 5 (12.2) | 36 (87.8) |  |  |  |
| **Currently in school** |  |  |  |  |  |
| Yes | 26 (25.2) | 77 (74.8) | 4.263 | 1 | **0.039** |
| No | 18 (14.4) | 107 (85.6) |  |  |  |
| **Highest educational level** |  |  |  |  |  |
| None | 1 (8.3) | 11 (91.7) | 8.537 | 4 | 0.074 |
| Primary | 1 (3.1) | 31 (96.9) |  |  |  |
| Secondary | 23 (21.5) | 84 (78.5) |  |  |  |
| Vocational | 6 (20.7) | 23 (79.3) |  |  |  |
| Tertiary | 13 (27.1) | 35 (72.9) |  |  |  |
| **Marital status** |  |  |  |  |  |
| Single | 13 (17.1) | 63 (82.9) | 8.306 | 3 | **0.040** |
| Married | 22 (16.9) | 108 (83.1) |  |  |  |
| Divorced/ Separated | 7 (46.7) | 8 (53.3) |  |  |  |
| Widowed | 2 (28.6) | 5 (71.4) |  |  |  |
| **Marriage type (if ever married)** |  |  |  |  |  |
| Monogamous | 19 (16.1) | 99 (83.9) | 5.988 | 1 | **0.014** |
| Polygamous | 12 (35.3) | 22 (64.7) |  |  |  |
| **Religion** |  |  |  |  |  |
| Christianity | 33 (18.2) | 148 (81.8) | 0.978 | 2 | 0.613 |
| Islam | 8 (25.8) | 23 (74.2) |  |  |  |
| Traditional | 3 (18.8) | 13 (81.2) |  |  |  |
| **Tribe** |  |  |  |  |  |
| Yoruba | 35 (18.2) | 157 (81.8) | 1.126 | 3 | 0.771 |
| Hausa | 2 (20.0) | 8 (80.0) |  |  |  |
| Igbo | 6 (27.3) | 16 (72.7) |  |  |  |
| Others | 1 (25.0) | 3 (75.0) |  |  |  |
| **Apprentice** |  |  |  |  |  |
| Yes | 7 (7.7) | 84 (92.3) | 13.098 | 1 | **<0.001** |
| No | 37 (27.0) | 100 (73.0) |  |  |  |
| **Years of experience** |  |  |  |  |  |
| <1 | 4 (6.7) | 56 (93.3) | 9.823 | 4 | **0.044** |
| 1-5 | 6 (17.6) | 28 (82.4) |  |  |  |
| 6-10 | 8 (22.2) | 28 (77.8) |  |  |  |

|  |  |  |
| --- | --- | --- |
| 11-20 | 5 (23.8) | 16 (76.2) |
| >20 | 21 (27.3) | 56 (72.7) |

CA who are within the age range 31-40 years is 3.4 times more likely to use tobacco than those ≤ 20 years (OR=3.410; CI=1.476-7.878) **(Table 4)**. Similarly, those that are currently in school and those who are divorced/ separated are twice likely (OR 2.007; CI= 1.026-3.927) and 4.2 times more likely (OR=4.240; CI=1.307-13.759) to use tobacco than those not in school and those single respectively. Furthermore, CA who were not apprentice were 4.4 times more likely to smoke than those who are apprentices (OR=4.440; CI=1.882-10.475). Moreover, CA have 6-10 years (OR=4.000; CI= 1.109-14.432), 11-20 years (OR=4.375; CI=1.050-18.234), and >20 years (OR=5.250; CI=1.693-16.278) of experience are four times, 4.3 times, and 5.2 times more likely to smoke than artisans with less than 1 year of experience respectively **(Table 4)**.

**Table 4:** Adjusted Odd Ratios for the associated factors of ever smokers among respondents.

| Variable | AOR | 95% CI for AOR |  |  |
| --- | --- | --- | --- | --- |
|  |  | Lower | Upper | p-value |
| Age group (in years) |  |  |  |  |
| ≤ 20 | 1.000 |  |  |  |
| 21 – 30 | 0.585 | 0.218 | 1.567 | 0.282 |
| 31 – 40 | 3.410 | **1.476** | **7.878** | **0.003** |
| > 40 | 0.673 | 0.222 | 2.042 | 0.482 |
| Currently in school |  |  |  |  |
| Yes | 2.007 | **1.029** | **3.927** | **0.039** |
| No | 1.000 |  |  |  |
| Marital status |  |  |  |  |
| Single | 1.000 |  |  |  |
| Married | 0.987 | 0.465 | 2.096 | 0.973 |
| Divorced/ Separated | 4.240 | **1.307** | **13.759** | **0.012** |
| Widowed | 1.939 | 0.339 | 11.101 | 0.451 |
| Marriage type (if ever married) |  |  |  |  |
| Monogamous | 1.000 |  |  |  |
| Polygamous | 2.842 | **1.205** | **6.701** | **0.040** |
| Apprentice |  |  |  |  |
| Yes | 1.000 |  |  |  |
| No | 4.440 | **1.882** | **10.475** | **<0.001** |
| Years of Experience |  |  |  |  |
| < 1 | 1.000 |  |  |  |
| 1 – 5 | 3.000 | 0.782 | 11.504 | 0.097 |
| 6 – 10 | 4.000 | **1.109** | **14.432** | **0.026** |
| 11 – 20 | 4.375 | **1.050** | **18.234** | **0.031** |
| > 20 | 5.250 | **1.693** | **16.278** | **0.002** |

**Figure 1:**
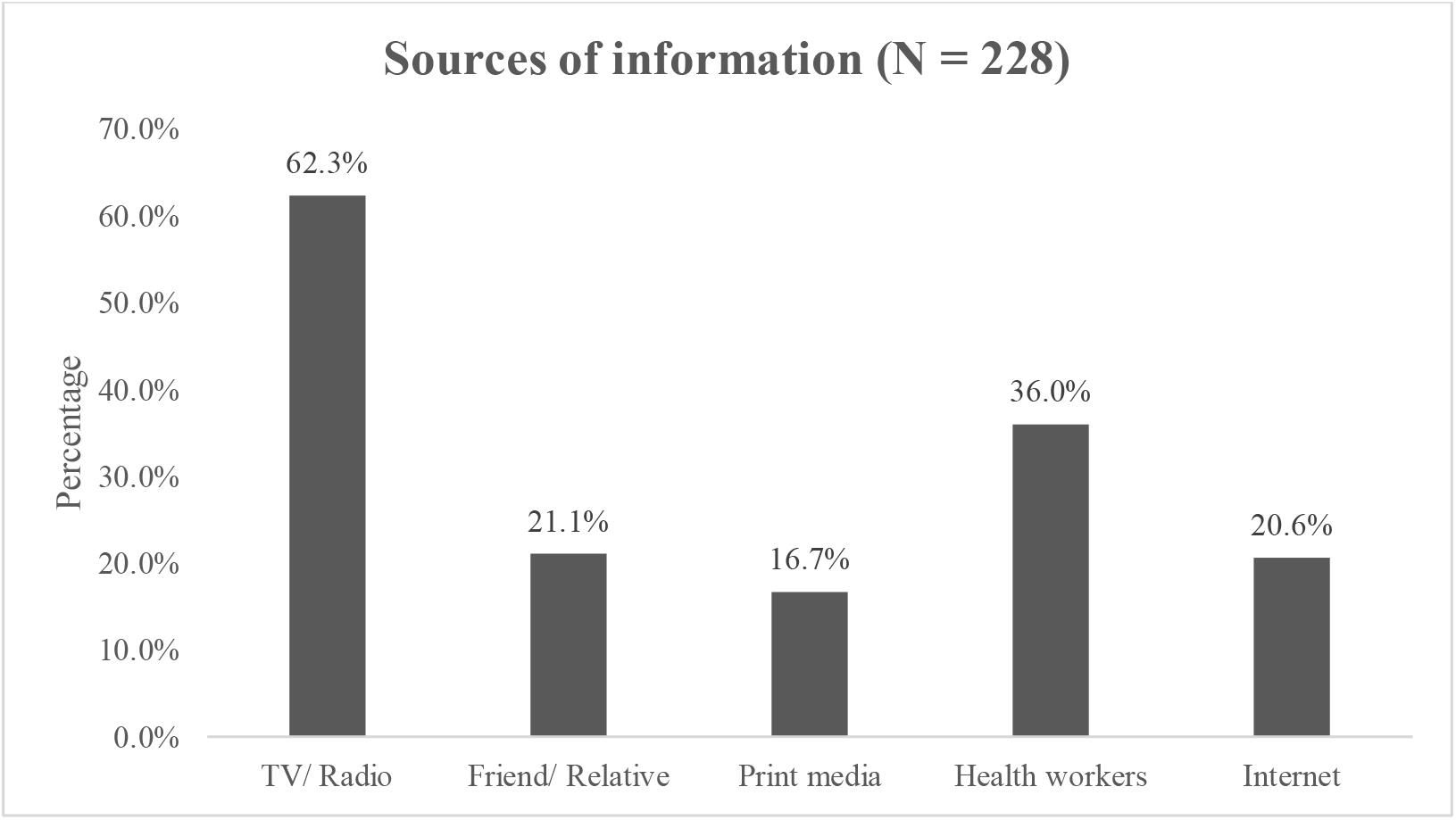
Respondents’ sources of information about tobacco-related health information. More than three-fifths (62.3%) of the respondents get health information on tobacco use from TV/radio, and print media was found to be the least explored means of getting health information. Also, amongst the respondents, 36% get health information from health workers while 21% and 20.6% get health information from friends /relatives and the internet, respectively.

**Figure 2:**
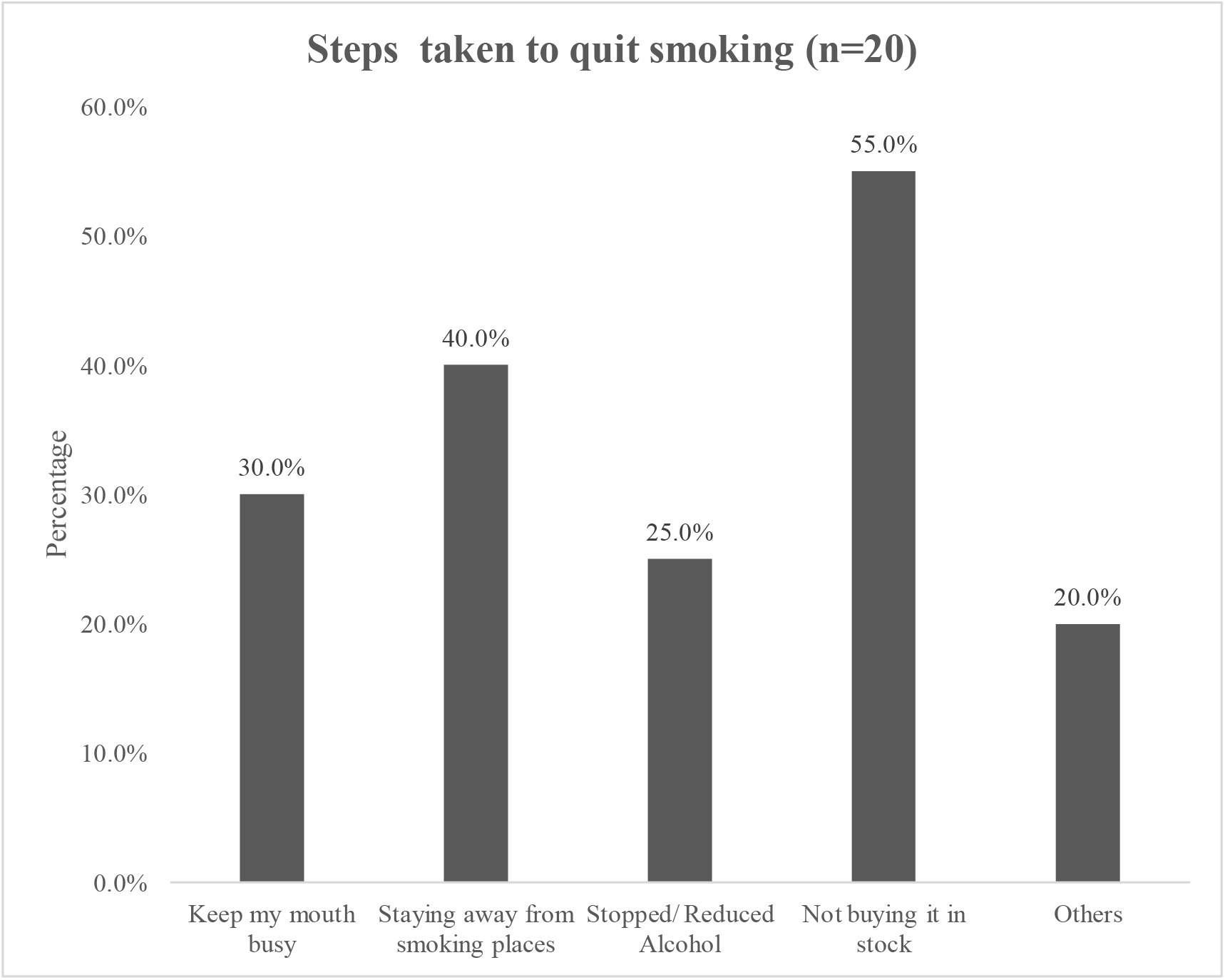
Steps were taken to quit smoking. More than half (55%) of the respondents tried quitting smoking by not buying cigarettes in stock, while a quarter (25%) had attempted quitting by stopping the intake of alcohol. Another 30% had attempted quitting by keeping their mouth busy with something else. Furthermore, when asked about smoking restriction at the worksite, almost two-thirds of respondents (64.9%) said there was no smoking restriction at work sites.

## DISCUSSION

In this study, the prevalence of ever used tobacco amongst CA was 19.3% and this was higher than the prevalence of ever smokers in the general population in Nigeria which was 3.9%.^9^ Similarly, a survey analysis of national data between 1992 - 2007 in the United States also gave the prevalence of ever smokers among construction artisans in the United States to be 38%^11^ and in Brazil, a cross-sectional survey among 418 male construction workers ever smoke prevalence was found to be 72.4%.^15^ In these countries, the prevalence of ever used tobacco in the general population was 30%^11,15^ and 56.9%^15^ in the United States and Brazil respectively. Therefore, the prevalence of ever used tobacco was particularly higher amongst this group of workers. However, the prevalence of ever smoke amongst CA in this study which was 19.3% is quite lower when compared to the prevalence of ever smoke amongst CA in Brazil which was 72.4%.^15^ This may not be unconnected to the fact that the prevalence of smoking in the general population in Brazil (56.9%)^15^ was far higher than the prevalence of ever smokers in the general population in Nigeria (3.9%).^3^ It must also be stressed that the prevalence of ever smokers in the general population in Nigeria was low possibly because some adults for cultural, spiritual, and social reasons would not admit having ever used tobacco.

In this study, several factors were identified to be associated with smoking. The age of artisans, marital status, and marriage type of CA, whether or not CA were presently in school, years of experience, and level of expertise of the CA were found to be significantly associated with tobacco use amongst CA. Artisans who are within the age range 31-40 years were 4.4 times more likely to smoke than those < 20 years. This was also in line with what Gavioli et al. (2014) found in a similar cross-sectional survey of 418 CA; CA who were more than 36 years of age have a high risk of tobacco use. Barbeau *et al*. (2004) and Okechechuwu *et al*. (2012) in their respective studies also highlighted a significant association between increasing age and smoking behavior among CA.^16,17^

Furthermore, CA who were divorced/separated were 4.2 times more likely to use tobacco. Liu et al. (2015) in a cross-sectional study amongst 5,380 migrants workers in China not only found 8.8 times increased risk of smoking in those who work in the construction sector but also found 2.2 times increased risk of smoking among these workers who were divorced.^18^ This could mean that physical stress of the construction work (which 31.8% of smokers admitted to) and smoking behavior of colleagues (which 86.4% of smokers admitted to) increases their uptake of smoking habit (Ajzen and Fishbein, 1980) on the one hand; and emotional and marital stress can increase their propensity to smoke on the other hand (Liu et al., 2015).^13,18^

Moreover, artisans with more than five years of work experience were more likely to be smoked when compared with those with lesser years of working experience. In this study, logistic regression analysis showed that CA with 6-10 years, 11-20 years, and > 20years of working experience were four times, five times, and six times respectively more likely to have ever smoker than those with less than one year experience. This was also in tandem with the findings of Gavioli et al. (2014) in a cross-sectional study of substance use amongst CA in Brazil^15^. They opined that CA with more than 10 years of working experience had an increased risk of ever smoking tobacco. It can then be concluded that the longer an artisan stays on the job, the higher his chances of taking up a smoking habit. This could be because he would be more exposed to the stress and strain of the job which may demand the use of a substance such as tobacco to relieve. Moreover, the more years an artisan spends on his job, the larger his circle of friends who are his co-workers and who might also be a smoker. With time, he might also take up the smoking habit just to conform to his co-workers.

It is also striking to note that almost two-thirds (64.9%) of the respondents said there was no smoking restriction at the worksite. Asfar et al. (2018) also noted a lack of smoking intervention that is adapted to the work environment of Hispanics/ Latinos working in the construction industries in the United States.^19^ Unfortunately, this lack of smoking restrictions at worksites may serve as an impetus to continue smoking. The use of tobacco cessation interventions at the worksite has shown to impact significantly on the smoking habits of CA.^10,19,20^ A mixed-method intervention (focus group and survey data) used amongst members of the Carpenters’ District Council in the United States showed that 65% of its 144 current smoking members were willing to quit smoking post-intervention.^10^ It was also noted that the majority responded positively to tailored messages and images on tobacco cessation.

### Limitations

The information on tobacco use was self-reported and this might lead to reporting bias, but self-reported estimates of tobacco use from other studies have indicated that this kind of information is still valid (Syamlal et al., 2014).^20^ More so, triangulating this study with qualitative data on the perception of CA on smoking would have given additional information on the smoking behavior of CA.

## CONCLUSION

This study assessed the prevalence of tobacco use amongst construction artisans in the communities in Ekiti State. It was found that the prevalence of cigarette smoking was higher amongst construction artisans than in the general population. Furthermore, the predisposing factors for the increased prevalence of this risky behavior amongst this group of workers were also assessed; age, marital status, and years of experience were strongly associated with indulgence in tobacco usage by CA.

The impacts of indulging in this behavior span beyond the CA themselves. There are medical, social, economic, and public health implications of this behavior to their immediate families, communities, and society at large. Therefore, individuals, communities, and governments at all levels must strive to mitigate any social and health inequalities linked with this unhealthy behavior for a better and healthier society.

## Data Availability

The datasets used and/or analyzed during the current study are available from the corresponding author on reasonable request

## DECLARATIONS

### Ethics approval

Research approval was obtained from the University of Liverpool ethics committee and the Ethics and Research Review Committee of the Federal Teaching Hospital, Ido Ekiti, Ekiti State, Nigeria.

### Consent for publication

Not applicable

### Authors’ Contributions

Conception/design of the study-COO, CMB, KAD; data collection-COO, ORO; data analysis and interpretation-COO, ORO; article drafting-COO, CMB; Critical revision of the article-COO, CMB, KAD; final approval of the version to be published-all authors

## Acknowledgment

Commonwealth Scholarship Commission (CSC) for granting me a full scholarship to pursue my MPH degree at the University of Liverpool, United Kingdom, and supporting me throughout the time I was writing this dissertation.

